# Toward Community Surveillance: Detecting Intact SARS-CoV-2 Using Exogeneous Oligonucleotide Labels

**DOI:** 10.1101/2021.03.23.21254201

**Authors:** Thomas R. Carey, Molly Kozminsky, Jennifer Hall, Valerie Vargas-Zapata, Kristina Geiger, Laurent Coscoy, Lydia L. Sohn

## Abstract

The persistence of the COVID-19 pandemic demands a dramatic increase in testing efficiency. Testing pooled samples for SARS-CoV-2 could meet this need; however, the sensitivity of RT-qPCR, the gold standard, significantly decreases with an increasing number of samples pooled. Here, we introduce DIVER, a method that quantifies intact virus and is robust to sample dilution. DIVER first tags viral particles with exogeneous oligonucleotides, then captures the tagged particles on ACE2-functionalized beads, and finally quantifies the oligonucleotide tags using qPCR. Using spike-presenting liposomes and Spike-pseudotyped lentivirus as SARS-CoV-2 models, we show that DIVER can detect 1×10^5^ liposomes and 100 pfu lentivirus and can successfully identify positive samples in pooling experiments. Overall, DIVER is well-positioned for efficient sample pooling and expanded community surveillance.

## Introduction

With well over 120 million confirmed cases and 2.7 million deaths worldwide to date^1^, the COVID-19 pandemic continues to have an enormous toll on human life. While optimism grows with the introduction of vaccines, there is still uncertainty as to whether these vaccines can prevent asymptomatic transmission^2,3^ and how long immunity lasts^4^. Additionally, the highly transmissible SARS-CoV-2 variants (e.g. B.1.351, B.1.1.7) that are quickly taking root in many countries are of great concern, as they have been shown to have increased resistance to neutralizing antibodies in laboratory studies^5,6^. Taken together, it is clear that widespread mass testing—up to 14 million tests per day^7,8^ in the United States alone—will be needed for the foreseeable future. Sample pooling, an efficient method to test large numbers of people, would save on healthcare resources, improve turn-around times on results, and ultimately provide the high testing capacity necessary to meet this demand^9,10^.

Sample pooling using the gold standard for SARS-CoV-2 detection, RT-qPCR, has been widely investigated. Innovative algorithms have been proposed that dynamically adjust pool size based on local disease prevalence^11^ or assign samples to pools of varying size to maximize efficiency using artificial intelligence^12^. However, sample pooling using RT-qPCR is inherently limited: because its sensitivity decreases as sample dilution increases, RT-qPCR’s false negative rate increases with sample pool size^9^. As an example, in four out of 24 pools of nine clinical SARS-CoV-2 samples, weak-positive specimens (those with a threshold cycle, or C_T_, between 31 and 35) were not detected, while these same specimens were detected in all pools of five samples^13^. Beyond RT-qPCR’s decreasing sensitivity with increasing pool size, it is susceptible to amplifying residual fragments of the viral genome, even in the absence of infectious virus^14-16^. Consequently, RT-qPCR is of limited use in assessing whether an individual is contagious^3^. Thus, there is an urgent need for novel assays that retain high sensitivity when samples are diluted for sample pooling and that detect only intact virus, which is better correlated with infectivity than viral RNA^14^.

We have developed an innovative, highly sensitive method, Detection of Intact Virus by Exogenous-nucleotide Reaction (DIVER), to detect and quantify SARS-CoV-2 in pooled samples for community surveillance. DIVER is based on: 1) labeling viral particles with exogenous single-stranded DNA oligonucleotides; 2) capturing the labeled viral particles with paramagnetic beads coated with ACE2, the primary receptor on human cells to which SARS-CoV-2 binds; and 3) performing qPCR on the *oligonucleotide labels*, which produces a signal that increases linearly with viral particle count (Fig. 1a). The oligonucleotide-labeling method in DIVER is adapted from one we previously applied to engineer cell-cell and cell-ligand interactions^17^ and to quantify tumor-derived extracellular vesicles (EVs) in biofluids^18^.

**Figure 1:**
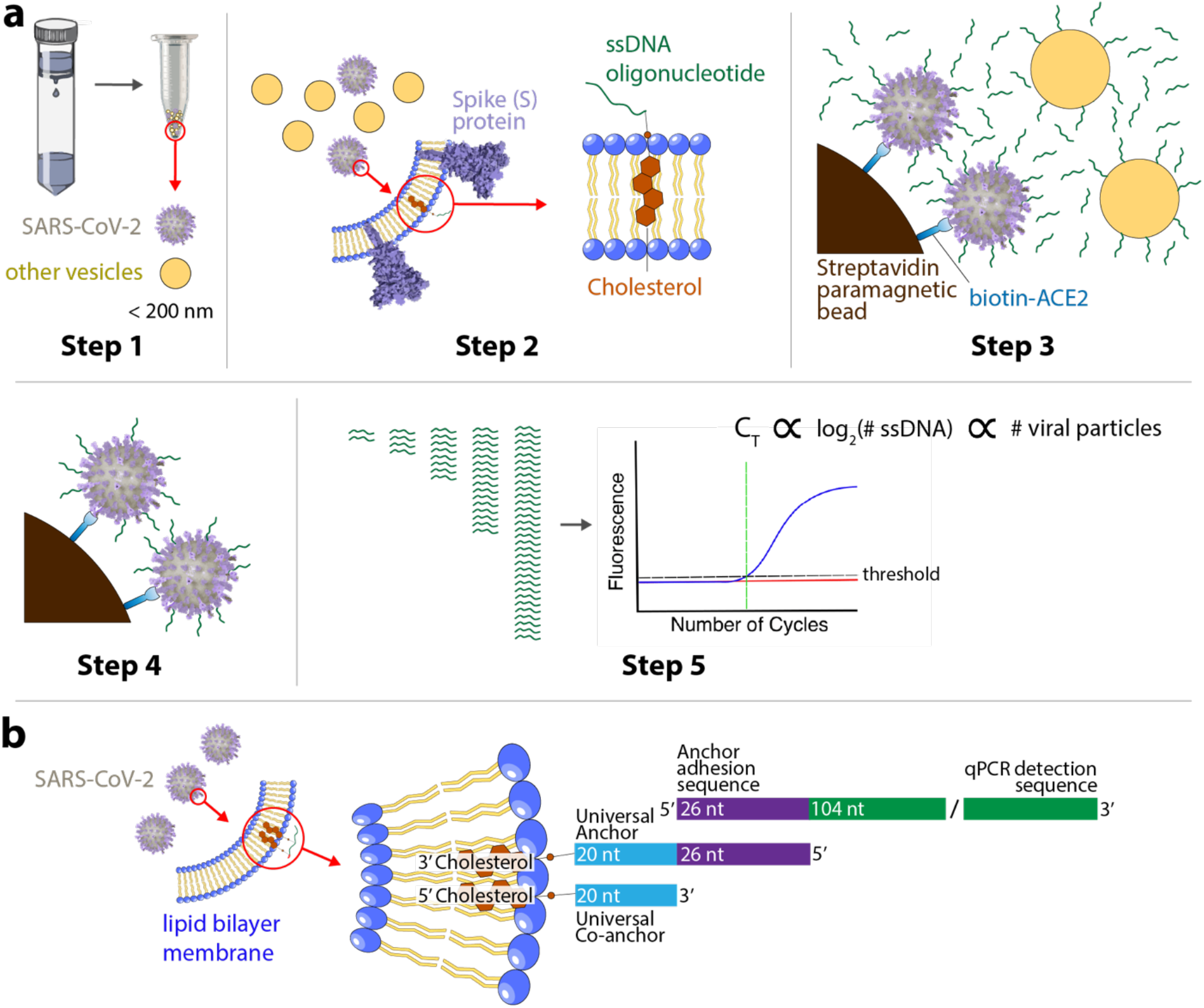
Oligonucleotide labeling of virus particles captured via spike-ACE2 interaction for quantitative detection. **a** Workflow for isolating, labeling, capturing, and quantifying nanoparticles displaying spike protein. Step 1: Isolate lipid bilayer nanoparticles < 200 nm in diameter using membrane affinity columns; Step 2: label isolated nanoparticles, including enveloped viruses and background extracellular vesicles (EVs), with cholesterol-tagged ssDNA oligonucleotides; Step 3: capture ssDNA oligonucleotide-labeled SARS-CoV-2, or models thereof, on ACE2-labeled paramagnetic beads; Step 4: wash to remove unbound oligonucleotides and background EVs; Step 5: using qPCR with primers specific to oligonucleotide labels, quantify ssDNA, which is directly correlated to the number of SARS-CoV-2 particles. **b** Details of oligonucleotide labeling scheme in Step 2 in (**a**). A 3’ cholesterol-tagged universal anchor oligonucleotide, prehybridized to a qPCR detection oligonucleotide, self-embeds in the lipid bilayer membrane. A 5’ cholesterol-tagged universal co-anchor oligonucleotide subsequently self-embeds into the lipid bilayer membrane and hybridizes with the first 20 nucleotides of the universal anchor, thereby stabilizing the complex^36^.

We utilized two models to demonstrate DIVER safely under Biosafety Level 2 conditions: liposomes displaying the SARS-CoV-2 spike protein^19^ and a Spike-pseudotyped lentivirus. With these models, we show that DIVER is uniquely suited for sample pooling, as its sensitivity is relatively unaffected by sample dilution. Our results demonstrate the promise of DIVER in expanding and improving community surveillance via sample pooling.

## Results

### Synthetic oligonucleotides label the lipid bilayer of SARS-CoV-2 model that is captured specifically on ACE2-functionalized beads

As SARS-CoV-2 is an enveloped virus, we first employed liposomes to verify labeling of lipid-bilayer nanoparticles with synthetic oligonucleotides and to capture specifically these particles onto beads via spike-ACE2 interaction. The liposome model we used displayed the S1 subunit of the SARS-CoV-2 spike protein, which contains the receptor binding domain (RBD), in an orientation that binds to ACE2^19^ (hereafter referred to as spike-liposomes) and were comparable in diameter (125.9 ± 27.3 nm; Fig. S1) to SARS-CoV-2.

We utilized oligonucleotide labels consisting of three components: a cholesterol-tagged universal anchor, a cholesterol-tagged co-anchor^20^, and a detection oligonucleotide that contains a 104 nt non-human/non-viral sequence that can be specifically amplified using qPCR (Fig. 1b and Methods). We verified successful oligonucleotide labeling of spike-liposomes by capturing, via hybridization, fluorescent spike-liposomes onto a surface functionalized with complementary oligonucleotides (Fig. S2). We also verified successful capture of spike-liposomes on ACE2-functionalized beads (ACE2 beads) by incubating a dilution series of fluorescent spike-liposomes (1×10^6^-1×10^9^ particles) with 1×10^6^ paramagnetic beads functionalized with either human ACE2 or anti-CD63 antibody (a negative control), and subsequently quantified the mean fluorescent intensity (Fig. S3). In contrast to anti-CD63 beads (aCD63 beads), for which we observed no significant correlation (r^2^ = 0.81, p = 0.09), the fluorescent intensity of ACE2 beads was strongly correlated with spike-liposome number (r^2^ > 0.99, p = 0.0024). These data strongly support that cholesterol-oligonucleotides self-embed in the membrane of lipid bilayer nanoparticles, consistent with previous findings^21^, and that ACE2 beads can readily sequester lipid bilayer nanoparticles displaying spike protein for subsequent qPCR-based detection.

### qPCR signal directly correlates with number of spike-liposomes in a heterogenous mixture

After successfully labeling spike-liposomes with oligonucleotides, we next demonstrated that the DIVER output signal—normalized qPCR threshold cycle—is correlated with the number of spike-liposomes. To quantify spike-liposomes, we used qPCR to amplify their oligonucleotides labels. Because DIVER concentrates oligonucleotide-labeled viral particles on ACE2 beads, it is relatively insensitive to sample volume (Fig. S4). Thus, we prepared spike-liposome samples in 10 µL PBS and analyzed these samples either directly or spiked into 10 µL samples containing either liposomes not displaying a surface protein (plain liposomes) or liposomes displaying human CD63 (CD63-liposomes). These samples were labeled with oligonucleotides and incubated with either ACE2 beads or aCD63 beads (see Methods).

We compared the qPCR signal from a spike-liposome dilution series (1×10^4^-1×10^7^ particles in 10 µL) with that from 1×10^7^ plain liposomes (Fig. 2a). We considered the difference in threshold cycle (dC_T_) between each sample and a no-liposome control containing all other assay components (i.e. oligonucleotide labels, ACE2 beads, and PCR master mix) processed in parallel to ensure that we quantified only oligonucleotide labels associated with liposomes. Samples with at least 1×10^5^ spike-liposomes had significantly higher dC_T_s than those containing just 1×10^7^ plain liposomes (1×10^5^, p < 0.01; ≥ 1×10^6^, p < 0.0001; Fig. 2a).

**Figure 2:**
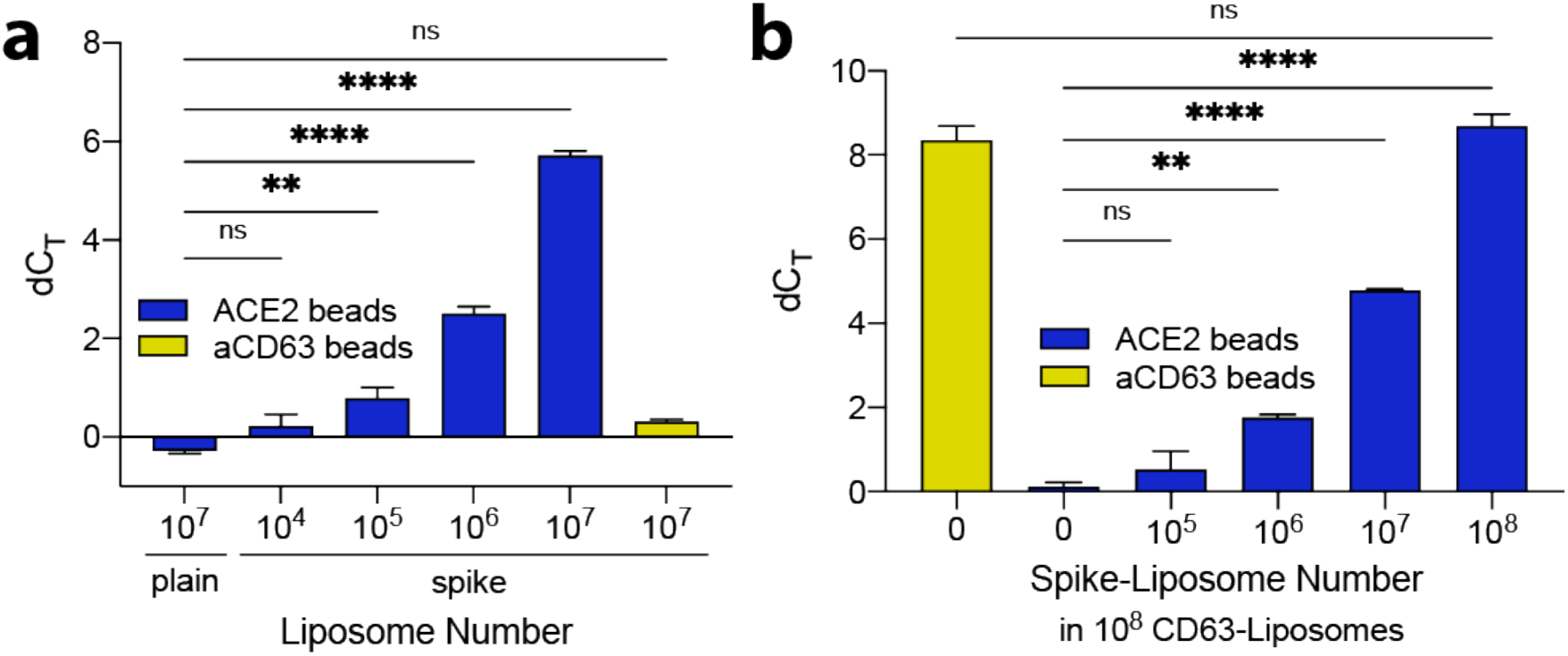
qPCR signal directly correlates with number of spike-liposomes in a sample. qPCR signal is presented as dC_T_, the difference in threshold cycle between each sample and a no-liposome control. **a** Dilution series of spike-liposomes suspended in buffer. Solutions containing ≥ 10^5^ spike-liposomes produced a signal significantly greater than 10^7^ plain liposomes when incubated with ACE2 beads. 10^7^ spike-liposomes produced a signal comparable to 10^7^ plain liposomes when incubated with aCD63 beads, indicating the qPCR signal results specifically from the capture of spike-liposomes onto ACE2 beads. **b** Dilution series of spike-liposomes in a background of 10^8^ CD63-liposomes. When incubated with ACE2 beads, solutions containing ≥ 10^6^ spike-liposomes produced a signal significantly greater than those containing just 10^8^ CD63-liposomes (0 spike-liposomes). As a control, 10^8^ CD63-liposomes with no spike-liposomes were incubated with aCD63 beads. The qPCR signal was comparable to that of 10^8^ spike-liposomes incubated with ACE2 beads, indicating that the capture of liposomes is highly specific. For all panels, error bars represent SEM; ns: not significant, *: p<0.05, **: p<0.01, ****: p<0.0001, one-way ANOVA with Dunnett’s (**a**) or Bonferroni’s (**b**) multiple comparisons test, with n = 3 independent replicates, each with n = 3 technical replicates. For detailed statistical information, see Table S1 (**a**) and Table S2 (**b**).

Biofluids contain abundant EVs (e.g., ∼10^8^/mL in saliva^22^) that are similar in size to SARS-CoV-2 and would also be labeled with oligonucleotides. To demonstrate DIVER’s specificity and ensure that labeling of off-target lipid bilayer nanoparticles, such as EVs, would not affect sensitivity, we used CD63-liposomes, as CD63 is a tetraspanin commonly present on the EV surface^23^. We found the difference in dC_T_ between samples containing at least 1×10^6^ spike-liposomes and 1×10^8^ CD63-liposomes or 1×10^8^ plain liposomes was significant (p < 0.01 and p < 0.0001; Fig. 2b and Fig. S5, respectively), suggesting that DIVER retains high sensitivity even in the presence of biologically relevant numbers of off-target lipid bilayer nanoparticles. To establish that ACE2 beads do not capture CD63-liposomes, we compared C_T_ values for 1×10^8^ CD63-liposomes and a no-liposome control (i.e. oligonucleotide labels and ACE2 beads added to PBS; 20 µL total volume). We observed no difference in C_T_ (p > 0.99; Fig. 2b), indicating that non-specific binding of CD63-liposomes on ACE2 beads was undetectable. Furthermore, we observed no difference in signal from 1×10^8^ CD63-liposomes captured on aCD63 beads and 1×10^8^ spike-liposomes (in a background of 1×10^8^ CD63-liposomes) captured on ACE2 beads (p = 0.92; Fig. 2b), verifying that the absence of signal for CD63-liposomes incubated with ACE2 beads was indeed a result of undetectable levels of nonspecific binding, not differences in oligonucleotide label densities between CD63-and spike-liposomes. To further illustrate our method’s specificity, we incubated 1×10^7^ spike-liposomes with ACE2 beads in a background of 10^5^-10^8^ CD63-liposomes (20 µL total volume). For each 10-fold increase in CD63-liposomes, dC_T_ decreased by 0.20 cycles (Fig. S6). These results establish that the signal we observe for spike-liposomes incubated with ACE2 beads is specifically due to amplification of spike-liposome-associated oligonucleotide labels.

### DIVER detects 100 pfu of novel SARS-CoV-2 Spike-pseudotyped lentivirus

We next validated DIVER’s ability to detect low numbers of viral particles displaying spike protein in a heterogenous mixture using an improved SARS-CoV-2 Spike-pseudotyped lentivirus that infects ACE2-expressing cells (293T-hACE2) more efficiently than the previously reported Spike-pseudotyped lentivirus^24^. The importance of the Spike-VSV-G chimera is that it results in biologically relevant virus:EV ratios in cell-culture media (approximately 1:400 ^22,25-27^).

To generate the Spike-VSV-G chimera, we mapped the domains of the SARS-CoV-2 Spike and VSV-G using existing literature^28^ and the Uniprot database, respectively. We designed this chimera to contain the transmembrane (aa 468-488) and cytoplasmic (aa 489-511) domains of VSV-G fused to the ectodomain of Spike (aa 1-1213) (Fig. 3a and Methods). The ectodomain of Spike includes both the receptor binding and fusion domains of this protein. We generated pseudotyped lentiviral particles with this Spike-VSV-G chimera and tested whether we could improve infection of target cells without compromising the specificity of Spike for the ACE2 receptor. Indeed, we observed a higher percentage of infected 293T-hACE2 cells using Spike-VSV-G in comparison to WT Spike (Fig. S7). Importantly, as expected, both pseudotypes poorly infected 293T cells lacking the ACE2 receptor. We calculated viral titers as previously described^24^ and found an average ∼2.5-fold increase in viral titers for Spike-VSV-G in comparison to WT Spike. Given these results, we chose to use the Spike-VSV-G chimera for DIVER experiments.

**Figure 3:**
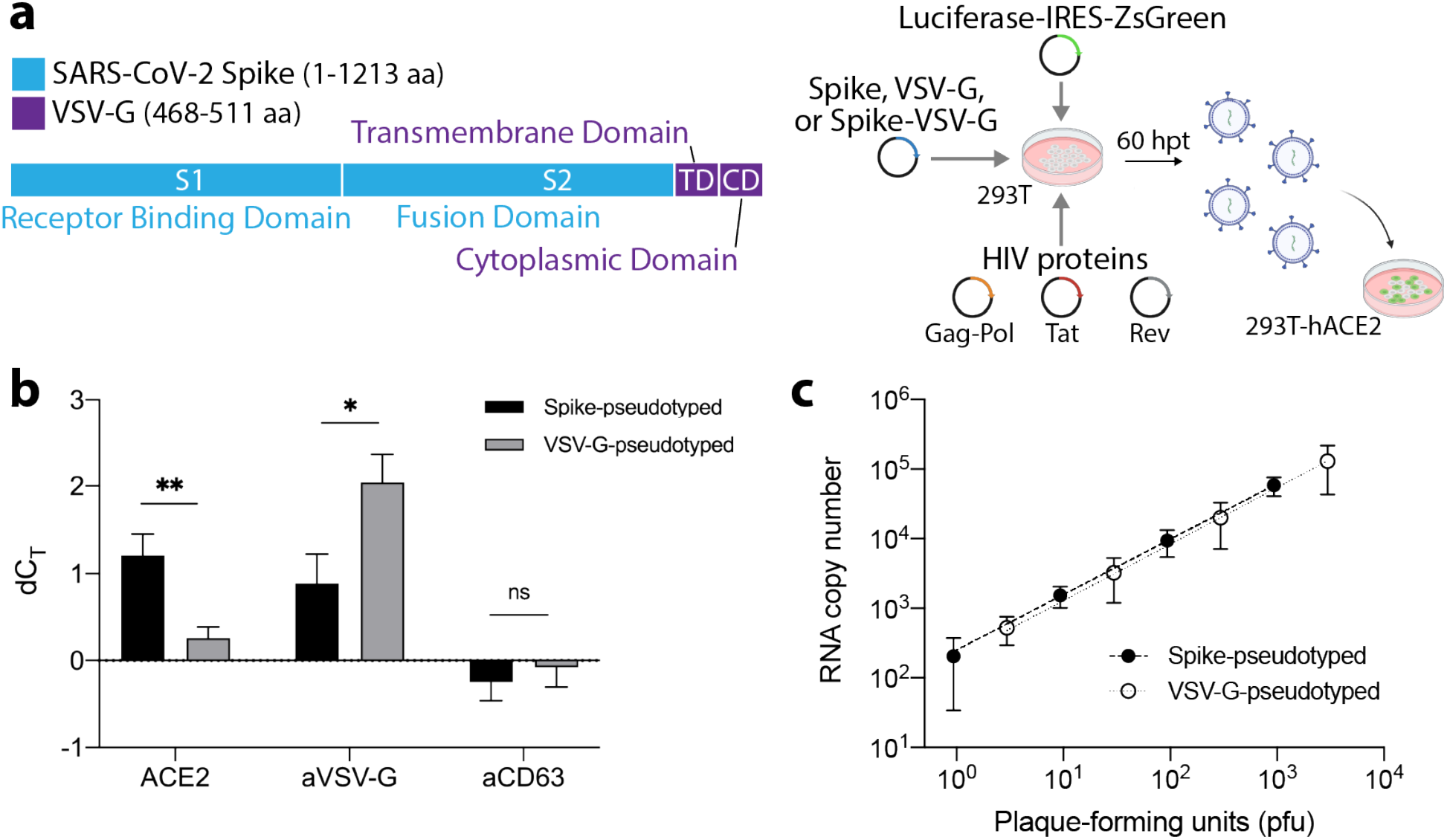
Design, production, and detection of 100 pfu SARS-CoV-2 Spike-pseudotyped lentivirus with DIVER. **a** Schematic showing the domain architecture of the SARS-CoV-2 Spike-VSV-G chimeric construct. This chimeric construct, in addition to plasmids encoding WT SARS-CoV-2 Spike and VSV-G, were used to generate pseudotyped lentivirus as previously described^24^. **b** qPCR signal, where dC_T_ is the difference in threshold cycle between lentivirus samples and no-virus control, was significantly greater for 100 pfu Spike-pseudotyped lentivirus than 100 pfu VSV-G-pseudotyped lentivirus captured onto ACE2 beads. Conversely, qPCR signal was significantly greater for 100 pfu VSV-G-pseudotyped lentivirus than that for 100 pfu Spike-pseudotyped lentivirus captured onto anti-VSV-G beads. aCD63 beads were employed to capture background EVs. Signals from Spike-and VSV-G-pseudotyped samples did not significantly differ from one another or from those of no-virus control samples, confirming that similar numbers of EVs were present in Spike- and VSV-G pseudotyped lentivirus and no-virus control cell culture supernatants. **: p<0.01, *: p<0.05, nested t-test (two-sided) to compare Spike-pseudotyped and VSV-G-pseudotyped lentivirus for each marker, n=4, each with n=3 technical replicates. For detailed statistical information, see Table S3. **c** RT-qPCR quantification of lentivirus indicates that 100 pfu Spike-pseudotyped lentivirus corresponds to an RNA copy number of approximately 9816, while 100 pfu VSV-G pseudotyped lentivirus corresponds to an RNA copy number of approximately 8365; n=2. For all panels, error bars represent SEM.

We analyzed 100 plaque-forming units (pfu) of Spike-pseudotyped lentivirus, 100 pfu of VSV-G-pseudotyped lentivirus, and an equivalent volume of no-virus control media, captured on either ACE2, anti-VSV-G (aVSV-G), or aCD63 beads. To ensure we quantified only oligonucleotide labels associated with lentiviral particles, we considered the dC_T_ between each sample and a no-virus control processed in parallel. The no-virus control was derived from cell culture supernatant from un-transfected 293T cells and contained all assay components (including oligonucleotide labels, ACE2 beads, and PCR master mix). We observed a significantly larger signal for Spike-pseudotyped lentivirus than for VSV-G-pseudotyped lentivirus incubated with ACE2 beads (p < 0.01) (Fig. 3b). We similarly observed a significantly larger signal for VSV-G-pseudotyped lentivirus than for Spike-pseudotyped lentivirus incubated with anti-VSV-G beads (p < 0.05). Because background EVs are likely to nonspecifically bind at low levels to ACE2 and aVSV-G beads, we verified that background EVs were present in similar numbers in Spike- and VSV-G pseudotyped lentivirus and no-virus control cell culture supernatants. Using DIVER with aCD63 beads to capture EVs, the qPCR signals from Spike- and VSV-G-pseudotyped samples did not significantly differ from one another or from signals from the no-virus control samples. These results demonstrate DIVER’s sensitivity and specificity to intact virus in a complex fluid containing EVs, proteins, and cell debris.

To compare DIVER against the current standard, RT-qPCR, for Spike-pseudotyped and VSV-G-pseudotyped lentiviruses, we reverse-transcribed and amplified a conserved region of the HIV-1 genome expressed in most lentiviruses^29^ (see Methods). By comparing threshold cycles to a standard curve that we constructed by serially diluting a control viral RNA template (Fig. S8), we obtained calculated infectivity coefficients of 98.2 RNA copies/pfu and 83.6 RNA copies/pfu for Spike and VSV-G-pseudotyped lentivirus, respectively (Fig. 3c). Thus, DIVER is able to detect robustly viral particles displaying spike protein corresponding to an RNA copy number of less than 10^4^.

### DIVER demonstrates high performance with large sample pools

While sample pooling enables a dramatic increase in the number of people who can be tested at once, there is a tradeoff in the decrease in sensitivity of RT-qPCR^9,30,31^. DIVER is uniquely suited for sample pooling because its output signal is more dependent on the *absolute number*, rather than the *concentration*, of labeled particles (Fig. S4). For our method, a 50x sample dilution results in a 7.7-fold (2.94 ± 0.15 cycle) decrease in signal vs. an 80-fold (5.42 ± 1.84 cycle) decrease in signal with RT-qPCR^30^. We demonstrated DIVER’s sample pooling capability by performing two different blinded experiments. We used three rounds of pooling (Fig. 4) to determine the positive sample, representing a 42% reduction in the number of tests required compared to processing each sample individually.

**Figure 4:**
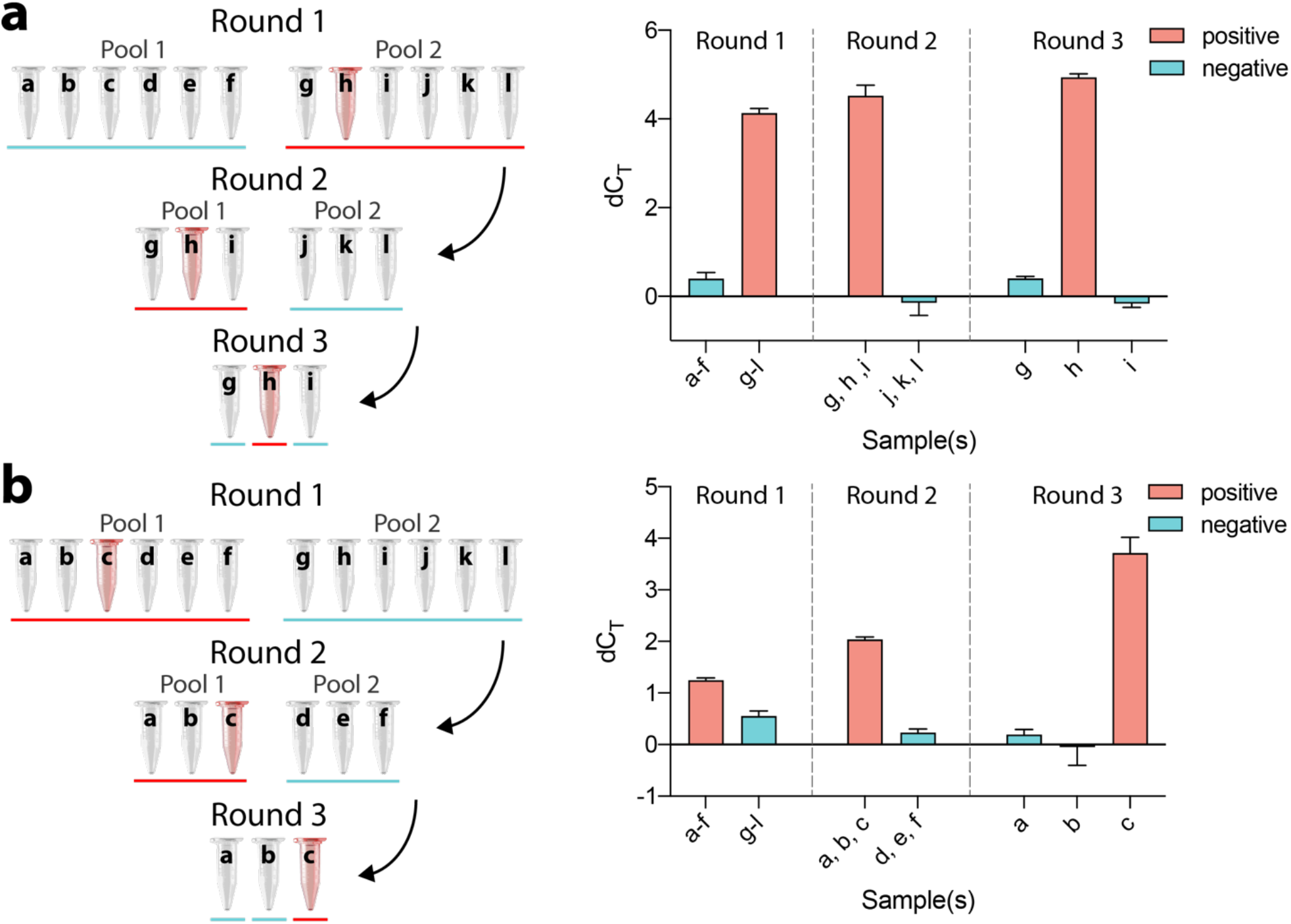
DIVER successfully detects positive samples in pooling experiments. Sample pooling enables a substantial reduction in resource usage for COVID-19 testing. In the pooling scheme shown here, the total number of qPCR tests is reduced from 12 to 7, a 42% reduction. A red bar indicates a pool or sample that tests positive, and a red tube indicates a positive sample. **a** Blinded demonstration of sample pooling using DIVER with liposomes. Twelve samples were prepared, each containing 10^7^ CD63-liposomes per µL; one sample also contained 10^6^ spike-liposomes per µL. In Round 1, 10 µL of each sample was combined into pools of six. In Round 2, for the pool that was identified as positive, 10 µL of each sample was combined into pools of three. Finally, in Round 3, 10 µL of each sample in the positive pool was analyzed individually. DIVER correctly identified sample *h* as containing 10^6^ spike-liposomes per µL. qPCR signal is presented here as dC_T_, the difference in threshold cycle between each pool or sample and an equal-volume no-liposome control. **b** Blinded demonstration of sample pooling using DIVER with Spike-pseudotyped lentivirus. Similar to the pooling demonstration with liposomes in (**a**), twelve samples were prepared, each of which contained 4.02×10^6^/µL cell-culture derived EVs, and one sample also contained 20 pfu/µL Spike-pseudotyped lentivirus. The same procedure described in (**a**) was used to identify correctly sample *c* as containing 20 pfu/µL lentivirus. Here, dC_T_ is the difference in threshold cycle between each pool or sample and an equal-volume no-virus control. n = 3 technical replicates; error bars represent SEM. All other samples were separately confirmed to be negative (Figs. S9 and S10).

In the first pooling experiment, 12 mock samples contained 1×10^7^ CD63-liposomes/µL, and one sample also contained 1×10^6^ spike-liposomes/µL. 10 µL of each sample was processed for each round of pooling. Using a threshold of 0.5 cycles, DIVER correctly identified the positive pool or sample for each round (Fig. 4a, Table S4). For confirmation, all samples were then processed individually (Fig. S9); the dC_T_ for negative pools or individual samples was 0.032 ± 0.273 (mean ± standard deviation), emphasizing our method’s negligible background signal for lipid bilayer nanoparticles not displaying spike protein.

In the second pooling experiment, we challenged DIVER to identify one positive sample from a large pool of samples with biologically relevant virus:EV ratios. Here, 12 mock samples contained 4.02×10^6^/µL cell-culture derived EVs, and one sample also contained 20 pfu/µL Spike-pseudotyped lentivirus. As before, 10 µL of each sample was processed for each round of pooling, and using a threshold of 0.6 cycles, DIVER correctly identified the positive pool or individual sample for each round (Fig. 4b, Fig. S10, Table S5).

## Discussion

Community surveillance for COVID-19 continues to be limited by insufficient testing capacity. While sample pooling would help enable the necessary improvements in testing efficiency, RT-qPCR’s false negative rate increases with sample pool size^9^. To enable larger sample pools, we present a novel virus detection method—DIVER—in which intact virus is labeled with cholesterol-tagged oligonucleotides, captured on ACE2 beads, and quantified by amplifying the oligonucleotide labels using qPCR. DIVER addresses several of the shortcomings of RT-qPCR. It detects only intact virus, which is more indicative of infectiousness than viral RNA^14^, and it is better suited for sample pooling, as it is more robust to sample dilution. In addition, DIVER does not require RNA extraction or reverse transcription, eliminating the need for reagents that have faced supply-chain challenges globally. As we have demonstrated, DIVER is highly specific, capable of detecting 10^5^ spike-liposomes or 100 pfu Spike-pseudotyped lentivirus against a background of CD63-liposomes or EVs, respectively.

In our experiments involving Spike-pseudotyped lentivirus, DIVER’s sensitivity was limited by the display of the common EV markers CD9, CD63, and CD81 on lentiviruses^32^. Sensitivity would be greatly improved when detecting SARS-CoV-2, which is assembled in the endoplasmic reticulum-Golgi intermediate compartment^33^ and thus does not display these markers^34^, by removing EVs using negative selection. This strategy would increase differential signal between positive samples and controls by approximately one cycle (Fig. S4). Background signal could be further reduced by mitigating nonspecific interactions between capture beads and free oligonucleotide labels. In addition, specific SARS-CoV-2 variants could be probed using validated antibodies against SARS-CoV-2 surface proteins in place of ACE2 as the capture moiety.

DIVER’s greater sensitivity to the absolute number, rather than the concentration, of viral particles in a sample is critical to the detection of SARS-CoV-2 in inherently dilute samples such as the pooled biosamples^31^ and wastewater^35^ that are currently screened for community surveillance. Furthermore, DIVER is adaptable to any enveloped virus. Should a novel enveloped viral pathogen emerge in the future, DIVER could be rapidly deployed: the viral genome does not need to be sequenced, primers do not need to be designed and validated, and only one component (the capture moiety) needs to be changed. By enabling larger sample pools while reducing the risk of false negatives, DIVER is uniquely positioned to contribute to the expanded community surveillance necessary to suppress COVID-19 and future pandemics.

## Methods

### Nanoparticle Tracking Analysis

Liposomes were diluted 1:1000 in 1X PBS (Gibco) and analyzed using nanoparticle tracking analysis on a Nanosight NS300 (Malvern Panalytical). Five technical replicates of 30s each were captured at a syringe pump speed of 50 µL/min and with the camera sensitivity set to ‘13,’ and analyzed with the detection threshold set to ‘3’ in the Nanosight software. Mean and standard deviation were determined by averaging measurements of three independent liposome samples and then applying a least-squares Gaussian fit.

### Oligonucleotide sequence design

The universal anchor sequence length was increased by 5 nt on the 5’ end relative to the sequence reported by Todhunter et al.^36^ in order to increase duplex length^37^ when hybridized to the detection oligonucleotide, which was also purchased from IDT. The detection oligonucleotide was based on the *Arabidopsis thaliana* FRK1 gene because of its lack of sequence homology with human and viral genes. Binding sites for primers reported in the literature^38^ (Table S1) were located at nucleotides 2232-2252 (forward) and 2352-2373 (reverse) of the *Arabidopsis thaliana* FLG22-induced receptor-like kinase 1 (FRK1) gene^38^; the detection oligonucleotide was then constructed from nucleotides 2230-2282 and 2323-2375 (5’-3’). Finally, a sequence complementary to the final 26 nt of the 5’ end of the universal anchor (adhesion sequence) was added to the 3’ end of the detection oligonucleotide for a total length of 130 nt (Table S6).

### Oligonucleotide preparation

Universal anchor and co-anchor sequences^36,39^, 46 and 20 nucleotides (nt) in length and modified with 3’ and 5’ cholesterol tags, respectively, and detection oligo, 130 nt in length (Table S1), were purchased from IDT. All primers were also purchased from IDT. Detection and universal anchor oligonucleotides were hybridized by combining equimolar quantities and incubating at 37°C for 15 minutes. This solution was then stored at −20°C for up to 6 months.

### Liposome fabrication

Liposome fabrication and labeling were performed as described^19^. Briefly, dioleoyl phosphatidyl choline, dioleoyl phosphatidyl ethanolamine, dioleoyl phosphatidyl glycerol acid, cholesterol, and 1,2-dioleoyl-sn-glycero-3-((N-(5-amino-1-carboxypentyl)iminodiacetic acid)succinyl) (nickel salt) (Avanti Polar Lipids) were combined in a 23.8 : 17.8 : 11.9 : 41.7 : 5.0 molar ratio and dissolved in chloroform (Millipore-Sigma). DiO (Thermo Fisher) was added at 1 mol% to lipid films to fabricate fluorescent liposomes. Once the chloroform was dried with dry nitrogen (N_2_) gas, the films were left to dry completely in a vacuum desiccator overnight. The films were rehydrated with Dulbecco’s PBS for 2 hrs at 37 C, then vortexed and bath sonicated prior to an overnight incubation at 4 C. The resulting suspension was sequentially extruded through 400 nm then 100 nm pore membranes (Avanti Polar Lipids) and then stored at 4 C until use, with a maximum storage time of 1 week. Liposome size and concentration were consistent across batches (Fig. S1).

For liposomes displaying a surface protein, liposomes were combined with either His-tagged S1 subunit of spike protein (Abcam) or His-tagged human CD63 (Sino Biological) at a 1:1000 molar ratio of liposome to protein. The resulting solution was diluted to a total volume of 100 µL in 0.2% bovine serum albumin (BSA, Millipore-Sigma) and incubated at room temperature for 2 hrs before storing at 4 C until use. Protein-labeled liposomes were used within 2 days.

### Oligonucleotide labeling and bead-based isolation of lipid-bilayer nanoparticles

M-270 Streptavidin Dynabeads (Invitrogen) were first washed in 0.2% BSA in PBS 2x and then resuspended in 0.2% BSA in PBS at a concentration of 1 µg/µL. Beads were then functionalized with biotin-ACE2 (Acro Biosystems), biotin-anti-CD63 (H5C6, BioLegend), or biotin-anti-VSV-G (polyclonal, Thermo Fisher Scientific) at ratios of 2.95 pg/µg beads, 20 ng/µg beads, and 20 ng/µg beads, respectively, by incubating for 30 min under gentle rotation. Following functionalization, beads were washed thoroughly 2x with 0.2% BSA and resuspended in 3% BSA in PBS at a concentration of 1 µg/µL.

10 µL or 20 µL of each sample of liposome or virus was combined with 500 fmol of detection oligonucleotide pre-hybridized to the universal anchor and incubated for 15 minutes. 500 fmol of co-anchor oligonucleotide was then added, followed by 10 µL of functionalized M270 Streptavidin Dynabeads. The sample was then incubated for one hour, washed 5x with 100 µL 0.2% BSA in PBS, and resuspended in 26 µL nuclease-free water.

### DNA-directed patterning

DNA was patterned onto the surface of glass slides, as previously described^17^. Briefly, an aldehyde-silane-coated glass slide (Applied Microarrays) was spin coated with a positive photoresist (S1813, Microchem; 3000 RPM for 30s) then heated for 90s on a 100°C hotplate. To expose regions of the slide to be patterned with DNA, resist-coated slides were exposed to UV light (365 nm, 9.5 mW/cm^2^) for 45s, then developed in MF321 (Microchem), and finally rinsed with 18 MΩ deionized (DI) water. A solution of 20 nt amine-terminated oligonucleotide in 50 mM sodium phosphate buffer was dropcast to coat the developed areas of the slide and heated in an oven at 85°C until the liquid had dried. To perform reductive amination, slides were soaked in 0.25% sodium borohydride (Sigma-Aldrich) in 1X PBS for 10 minutes with light shaking prior to a DI water wash. Photoresist was removed with acetone (Gallade Chemical) and an additional DI water wash was performed. Slides were dried with N_2_ gas. DNA patterns were enclosed using polydimethylsiloxane (PDMS) flow cells, where the edges of the channel were sealed using PDMS stamping.

To create fluorescent patterns, liposomes were labeled with cholesterol oligonucleotides as described above. PDMS flow cells were blocked with 2% bovine serum albumin (BSA, Millipore-Sigma) for at least 30 minutes after which oligonucleotide-labeled liposomes were cycled through the flow cells 10 times to allow the oligonucleotide labels to hybridize with complementary oligonucleotides immobilized on the surface. Patterned liposomes were imaged using a FITC filter and 20x objective on an ImageXpress Micro High-Content Imaging System.

### Fluorescent liposome capture assay

M-270 Streptavidin Dynabeads (Invitrogen) were prepared and functionalized as described above. Serial dilutions of DiO-labeled liposomes displaying spike protein (spike-liposomes) were incubated for 2 hr with 1 µg beads functionalized with either biotin-ACE2 (ACROBiosystems) or biotin-anti-CD63 (BioLegend). After washing 3x with 0.2% BSA in PBS, samples were resuspended in 20 µL PBS. 5 µL of each sample was deposited on a microscope slide and covered with a coverslip. Beads were allowed to settle and then were imaged using an inverted fluorescent microscope (Nikon TE2000-E) at 60x magnification with a FITC filter. Mean fluorescent intensities of beads in each image were determined using the ‘Analyze Particles’ tool in ImageJ (NIH).

### qPCR

Samples suspended in nuclease-free water were mixed with qPCR master mix (Applied Biosystems PowerUp SYBR Green) and primers (Table S1; 50 nM final concentration; IDT). 20 µL reactions were added in triplicate to each PCR plate. qPCR parameters were programmed according to the Applied Biosystems PowerUp SYBR Green protocol. Briefly, samples were denatured at 95 C for 15 sec and annealed and extended at 60 C for 1 minute for 40 cycles. qPCR was run on a StepOne qPCR machine (Applied Biosystems). On each plate, a sample consisting of 2.5 × 10^6^ copies of FRK1 oligonucleotide was included; during analysis, the threshold was adjusted such that the threshold cycle for this standard was 27.00, thereby enabling comparison of data across PCR plates. Analysis was completed using StepOne Software v2.2.2. Oligonucleotide associated with liposomes or virus was differentiated from residual oligonucleotides on beads by subtracting the threshold cycle of a no-liposome or no-virus control, processed in parallel, from the threshold cycle of each sample (dC_T_).

### Generation of Spike-VSV-G chimera vector

The spike-VSV-G chimeric fragment was generated by SOEing PCR. The HDM-IDTSpike-fixK^24^ and pCMV-VSV-G (Addgene plasmid #8454) vectors were used to generate the chimeric insert. The Spike-VSV-G fragment was then cloned into HDM-IDTSpike-fixK using EcoRV and XhoI restriction enzymes. The final plasmid, HDM_IDTSpike-VSV-G fusion, was verified by sequencing.

### Cell culture for lentivirus production

293T cells were obtained from the UC Berkeley Cell Culture Facility. These cells were grown in DMEM (Gibco, 11995073) supplemented with 5% FBS (VWR, 89510-186) and 1X Penicillin-Streptomycin (100 U Penicillin, 100 µg/mL Streptomycin final concentrations; Genesee Scientific, 25-512). 293T-hACE2 cells were obtained from Jesse D. Bloom’s lab at the Fred Hutchinson Cancer Research Center and cultured in DMEM supplemented with 10% FBS and 1X Penicillin-Streptomycin. Both cell lines were cultured at 37 °C with 5% CO2.

### Lentivirus production

Production and titering of lentivirus were accomplished using the lentivirus pseudotyping strategy published by Crawford *et al*. (Sections 4.3 and 4.4)^24^. Briefly, 293T cells were seeded at 5×10^5^ cells/well in a 6-well plate. After 24 hrs, cells were transfected using Fugene HD (following manufacturer’s instructions; Promega) with the lentiviral transfer plasmid, packaging plasmids, and envelope plasmid using the Luciferase-IRES-ZsGreen as the lentiviral backbone and Spike, Spike-VSV-G, or VSV-G as the entry proteins. All plasmids used for the pseudotyping system were obtained from Jesse D. Bloom’s lab at the Fred Hutchinson Cancer Research Center. After 18 hours, fresh media (10% FBS in DMEM) was added slowly to the wells. Viral supernatant was harvested 60 hours post transfection, filtered through a 0.45 mm filter, and frozen at −80C before titering.

### Functional titering of lentivirus

Titering was performed with the following modifications to the protocol in Crawford *et al*. (Section 4.4)^24^. A tissue-culture-treated 96-well plate with no additional poly-L-lysine coating was used. In addition to 293T-hACE2 cells, 293T were used as control target cells. Undiluted and 1:3 dilutions of the lentiviral supernatants containing a final concentration of 10 µg/mL polybrene (Santa Cruz Biotechnology) were used. Cells were spinfected for 2 hr at 750 x g. After spinfection, the media was changed and the infection was allowed to proceed for 48 hrs. Cells were harvested with 0.05% Trypsin-EDTA (Gibco) and resuspended in staining buffer containing 1X PBS pH 7.4 (Gibco), 3% Fetal Bovine Serum (FBS) (VWR), and 1 mM EDTA (Fisher Scientific). Cells were analyzed by flow cytometry (Becton Dickinson LSRFortessa) using the FITC channel to detect ZsGreen and the data analyzed in FlowJo (v.10). Viral titers were calculated using the Poisson formula as described by Crawford *et al*.^24^.

### Purification of lentivirus from cell-culture supernatant for oligonucleotide-labeling assay

Frozen cell-culture supernatant containing lentivirus and extracellular vesicles (EVs) was thawed at 37 C and centrifuged at 5000 rcf for 5 min. Particles with a lipid bilayer membrane, i.e. lentivirus and EVs, were then purified using ExoEasy Maxi membrane affinity columns (Qiagen) per the manufacturer’s protocol. 400 µL eluate containing lentivirus and EVs in Buffer XE (Qiagen) was buffer exchanged into 1X PBS (Gibco) and concentrated to 62 µL using Amicon Ultra centrifugal filters with a nominal molecular weight limit of 10 kDa (Millipore Sigma).

### RT-qPCR lentivirus titration

Frozen cell-culture supernatant containing lentivirus was thawed at 37 °C. RNA was isolated from 150 µL cell-culture supernatant using a Nucleospin RNA Virus kit (Takara Bio) per manufacturer’s protocol. Isolated RNA suspended in 50 µL nuclease-free H_2_O was subsequently stored at −80 C. Serial dilutions of lentiviral RNA and RNA control template were processed with RT-qPCR using the Lenti-X qRT-PCR Titration Kit (Takara Bio) per manufacturer’s protocol. Briefly, triplicate 25 µL reactions were prepared containing either lentiviral RNA or RNA control template, Quant-X buffer, primers (200 nM final concentration), ROX reference dye LSR, Quan-X enzyme, and RT enzyme mix. One-step RT-PCR cycling parameters were as follows: reverse transcription by heating samples to 42 C for 5 min then 95 C for 10 sec, then 40 cycles of amplification by denaturing samples at 95 C for 5 sec, followed by annealing and extending at 60 C for 30 sec. Because virus was produced from stably transfected cell lines, a DNase I treatment was not performed. RT-qPCR was run on a StepOne qPCR machine (Applied Biosystems).

To calculate standard curves for each experiment, extreme outliers between technical replicates (individual wells containing identical samples) within each experiment were first removed. Extreme outliers were defined as wells for which the threshold cycle deviated by > 3 (standard deviations) from the median threshold cycle for that sample. A semi-log least-squares fit was then computed and used to calculate RNA copy number as a function of threshold cycle for lentivirus samples.

For each lentivirus sample, calculated quantities for six total technical replicates across two independent experiments were combined. Due to high variability between technical replicates, extreme outliers (calculated quantities that were > 3 from the median) were removed. Log-log least-squares fits were then computed for each lentivirus pseudotype to determine the approximate threshold cycle corresponding to a specific functional titer of each lentivirus. Using the standard curve, threshold cycles were converted to RNA copy number. The infectivity coefficient for each lentivirus pseudotype was then calculated as the ratio of RNA copy number to functional titer.

### Statistical analysis

All data are reported as the mean ± standard error of the mean. All statistical analysis and plotting were performed with GraphPad Prism 9.0.2. When comparing samples against a control, differences were analyzed using one-way ANOVA with Dunnett’s multiple comparison test (Figs. 2a and S5; Tables S1 and S8). When comparing specific samples pairwise, differences were analyzed using one-way ANOVA with Bonferroni’s multiple comparison test (Fig. 2b; Table S2). Differences between two groups for different markers were analyzed using a nested two-sided t-test (Fig. 3b; Table S3). For all comparisons, differences with p < 0.05 were considered significant. r^2^ values reported are Pearson’s Correlation coefficients (Fig. S3; Table S7).

## Data Availability

All data needed to evaluate the conclusions in this paper are available in the manuscript or supplementary materials.

## Acknowledgements

We thank N. Liu and S. Kitayama for experimental assistance, the J. Bloom Lab (Fred Hutchinson Cancer Research Center) and P.S. Kim Laboratory (Stanford University) for the 293T-hACE2 cells and lentiviral plasmids, the R. Schekman Lab for the use of their Nanoparticle Tracking Analysis instrument, UC Berkeley QB3 Cell and Tissue Analysis Facility, and O. Scheideler and S. Hinz for critical reading of this manuscript.

## Author contributions

Conceptualization: T.R.C., M.K., and L.L.S. Methodology: T.R.C., M.K., V.V.-Z., and L.L.S. Formal analysis: T.R.C., M.K., and V.V.-Z. Investigation: T.R.C., M.K., J.H., V.V.-Z., K.G. Resources: T.R.C., L.L.S., L.C. Visualization: T.R.C., M.K., and V.V.-Z. Supervision: L.L.S. Funding acquisition: L.L.S. and L.C. Writing – original draft: T.R.C., M.K., V.V.-Z., and K.G. Writing – review and editing: T.R.C., M.K., L.L.S., J.H., V.V.-Z., K.G., and L.C.

