## Supplementary Information for "Toward Community Surveillance: Detecting Intact SARS-CoV-2 Using Exogeneous Oligonucleotide Labels"

### Table S1: Statistics corresponding to Fig. 2a.

| | Mean 1 | Mean 2 | Mean Diff. | 95.00% CI of diff. | Summary | Adjusted P Value | q | DF |
|---|---|---|---|---|---|---|---|---|
| **$10^7$ CD63 vs. $10^7$ plain** | 0.314 | -0.282 | 0.596 | -0.1303 to 1.323 | ns | 0.1417 | 2.213 | 35 |
| **$10^4$ vs. $10^7$ plain** | 0.225 | -0.282 | 0.507 | -0.2191 to 1.234 | ns | 0.2625 | 1.884 | 35 |
| **$10^5$ vs. $10^7$ plain** | 0.788 | -0.282 | 1.070 | 0.3436 to 1.796 | ** | 0.0018 | 3.973 | 35 |
| **$10^6$ vs. $10^7$ plain** | 2.510 | -0.282 | 2.792 | 2.065 to 3.518 | **** | <0.0001 | 10.37 | 35 |
| **$10^7$ vs. $10^7$ plain** | 5.717 | -0.282 | 6.000 | 5.273 to 6.726 | **** | <0.0001 | 22.28 | 35 |

Dunnett's multiple comparisons test
alpha = 0.05

**Table S2: Statistics corresponding to Fig. 2b.** All conditions included a background of $10^8$ CD63 liposomes.

| | Mean 1 | Mean 2 | Mean Diff. | 95.00% CI of diff. | Summary | Adjusted P Value | t | DF |
|---|---|---|---|---|---|---|---|---|
| **$10^5$ vs. 0** | 0.532 | 0.124 | 0.409 | -0.6538 to 1.471 | ns | >0.9999 | 1.195 | 11 |
| **$10^6$ vs. 0** | 1.757 | 0.124 | 1.633 | 0.5707 to 2.696 | ** | 0.0029 | 4.774 | 11 |
| **$10^7$ vs. 0** | 4.781 | 0.124 | 4.658 | 3.595 to 5.720 | **** | <0.0001 | 13.62 | 11 |
| **$10^8$ vs. 0** | 8.690 | 0.124 | 8.566 | 7.504 to 9.629 | **** | <0.0001 | 25.04 | 11 |
| **$10^8$ ACE2 vs. CD63** | 8.690 | 8.353 | 0.337 | -0.8511 to 1.525 | ns | >0.9999 | 0.8805 | 11 |

Bonferroni's multiple comparisons test
alpha = 0.05

### Table S3: Statistics corresponding to Fig. 3b.

| Sample | ACE2 | aVSV-G |
|---|---|---|
| **P value** | 0.0051 | 0.0481 |
| **P value summary** | ** | * |
| **One- or two-tailed P value?** | Two-tailed | Two-tailed |
| **t, df** | t=4.294, df=6 | t=2.475, df=6 |
| **F, DFn, Dfd** | 18.44, 1, 6 | 6.125, 1, 6 |
| **Mean of column Spike** | 1.208 | 0.8885 |
| **Mean of column VSV-G** | 0.03922 | 2.04 |
| **Difference between means (Spike - VSV-G) ± SEM** | 1.168 ± 0.2721 | -1.152 ± 0.4654 |
| **95% confidence interval** | 0.5027 to 1.834 | -2.291 to -0.01306 |

**Table S4**: **Descriptive statistics for each pool for spike-liposome sample pooling**. Positive samples or pools are highlighted in gray.

| | | n | Mean | Std. Dev. | S.E.M. | Lower 95% CI | Upper 95% CI |
|---|---|---|---|---|---|---|---|
| **6 samples/pool** | a-f | 3 | 0.396 | 0.241 | 0.139 | -0.203 | 0.996 |
| | g-l | 3 | 4.130 | 0.186 | 0.107 | 3.670 | 4.590 |
| **3 samples/pool** | a, b, c | 3 | -0.103 | 0.215 | 0.124 | -0.637 | 0.432 |
| | d, e, f | 3 | 0.312 | 0.218 | 0.126 | -0.229 | 0.853 |
| | g, h, i | 3 | 4.520 | 0.410 | 0.236 | 3.510 | 5.540 |
| | j, k, l | 3 | -0.146 | 0.499 | 0.288 | -1.390 | 1.090 |
| **Individual samples** | a | 3 | 0.188 | 0.169 | 0.098 | -0.233 | 0.608 |
| | b | 3 | 0.193 | 0.049 | 0.028 | 0.071 | 0.315 |
| | c | 3 | -0.365 | 0.059 | 0.034 | -0.510 | -0.220 |
| | d | 3 | 0.167 | 0.469 | 0.271 | -0.997 | 1.330 |
| | e | 3 | -0.126 | 0.124 | 0.071 | -0.433 | 0.181 |
| | f | 3 | -0.431 | 0.084 | 0.048 | -0.639 | -0.223 |
| | g | 2 | 0.405 | 0.066 | 0.047 | -0.189 | 0.999 |
| | h | 3 | 4.930 | 0.146 | 0.084 | 4.570 | 5.300 |
| | i | 3 | -0.163 | 0.153 | 0.088 | -0.542 | 0.217 |
| | j | 2 | -0.170 | 0.055 | 0.039 | -0.665 | 0.324 |
| | k | 3 | -0.023 | 0.134 | 0.077 | -0.355 | 0.310 |
| | l | 3 | 0.344 | 0.350 | 0.202 | -0.525 | 1.210 |

**Table S5**: **Descriptive statistics for each pool for Spike-pseudotyped lentivirus sample pooling.** Positive samples or pools are highlighted in gray.

| | | n | Mean | Std. Dev. | S.E.M. | Lower 95% CI | Upper 95% CI |
|---|---|---|---|---|---|---|---|
| **6 samples/pool** | a-f | 3 | 1.250 | 0.080 | 0.046 | 1.050 | 1.450 |
| | g-l | 3 | 0.552 | 0.172 | 0.100 | 0.124 | 0.980 |
| **3 samples/pool** | a, b, c | 3 | 2.040 | 0.088 | 0.051 | 1.820 | 2.260 |
| | d, e, f | 3 | 0.232 | 0.126 | 0.073 | -0.082 | 0.546 |
| | g, h, i | 3 | 0.516 | 0.102 | 0.059 | 0.263 | 0.770 |
| | j, k, l | 3 | 0.248 | 0.376 | 0.217 | -0.685 | 1.180 |
| **Individual samples** | a | 3 | 0.192 | 0.172 | 0.099 | -0.234 | 0.618 |
| | b | 6 | -0.047 | 0.868 | 0.354 | -0.958 | 0.864 |
| | c | 3 | 3.710 | 0.523 | 0.302 | 2.420 | 5.010 |
| | d | 3 | 0.454 | 0.184 | 0.106 | -0.003 | 0.910 |
| | e | 3 | 0.445 | 0.203 | 0.117 | -0.060 | 0.950 |
| | f | 3 | 0.076 | 0.278 | 0.160 | -0.613 | 0.765 |
| | g | 3 | 0.433 | 0.388 | 0.224 | -0.530 | 1.400 |
| | h | 3 | 0.430 | 0.188 | 0.109 | -0.037 | 0.898 |
| | i | 3 | -0.229 | 0.299 | 0.172 | -0.971 | 0.513 |
| | j | 3 | -0.455 | 0.366 | 0.211 | -1.360 | 0.455 |
| | k | 3 | -0.269 | 0.246 | 0.142 | -0.880 | 0.343 |
| | l | 3 | 0.001 | 0.216 | 0.124 | -0.534 | 0.537 |

**Table S6**: **Oligonucleotide and primer sequences.** Name indicates distinct sequences, and colors indicate complementary regions that hybridize in our assay.

| Name | Modification | Sequence (5'-3') | Annotation |
|---|---|---|---|
| Universal Anchor | 3' Cholesterol-TEG | TGGAATTCTCGGGTGCCAAGGGAAT<br>TC GTAACGATCCAGCTGTCACT | Detection adhesion sequence<br>Co-anchor adhesion sequence |
| Universal Co-Anchor | 5' Cholesterol-TEG | AGTGACAGCTGGATCGTTAC | Anchor adhesion sequence |
| Detection Oligo | N/A | GAATTCCCTTGGCACCCGAGAATTCCA<br>TGAAGGAAGCGGTCAGATTTCAACA<br>GTTGTCGCTGGATCCATCGGTTGTTC<br>TTCTTGAAGTGATTACAGGCCAACCT<br>GCTATTCAGTCAGTCAGTCAGTCAGT | Anchor adhesion sequence<br>Detection sequence |
| FRK1 Forward Primer | N/A | CGGTCAGATTTCAACAGTTGTC | |
| FRK1 Reverse Primer | N/A | AATAGCAGGTTGGCCTGTAATC | |

**Table S7: Statistics corresponding to Fig. S3.**

| | ACE2 Beads | aCD63 Beads |
|---|---|---|
| **Pearson r** | 0.9976 | 0.9027 |
| **95% confidence interval** | 0.8879 to 1.000 | -0.4410 to 0.9980 |
| **R squared** | 0.9953 | 0.8148 |
| **P (two-tailed)** | 0.0024 | 0.0973 |
| **P value summary** | ** | ns |

Fluorescence vs. Liposome Number Correlation
alpha = 0.05

**Table S8: Statistics corresponding to Fig. S5.**

| | Mean 1 | Mean 2 | Mean Diff. | 95.00% CI of diff. | Summary | Adjusted P Value | q | DF |
|---|---|---|---|---|---|---|---|---|
| **0 vs. $10^5$** | 0.021 | 0.575 | -0.554 | -1.386 to 0.2781 | ns | 0.2685 | 1.736 | 25 |
| **0 vs. $10^6$** | 0.021 | 1.992 | -1.971 | -2.803 to -1.139 | **** | <0.0001 | 6.175 | 25 |
| **0 vs. $10^7$** | 0.021 | 5.063 | -5.042 | -5.874 to -4.210 | **** | <0.0001 | 15.8 | 25 |
| **0 vs. $10^8$** | 0.021 | 8.986 | -8.965 | -9.797 to -8.133 | **** | <0.0001 | 28.09 | 25 |

Dunnett's multiple comparisons test
alpha = 0.05

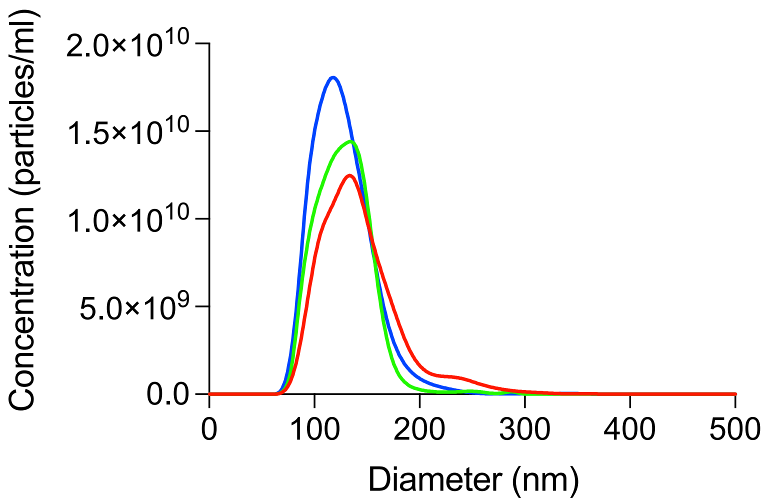


**Figure S1**: **Nanoparticle tracking analysis measurements of three independently fabricated batches of liposomes show highly reproducible size distribution and concentration.** Each colored line represents the average of five technical replicate measurements of an independent batch of liposomes. Liposomes were 125.9 ± 27.3 nm in diameter and were comparable in size to SARS-CoV-2, which are 60-140 nm in diameter[40].

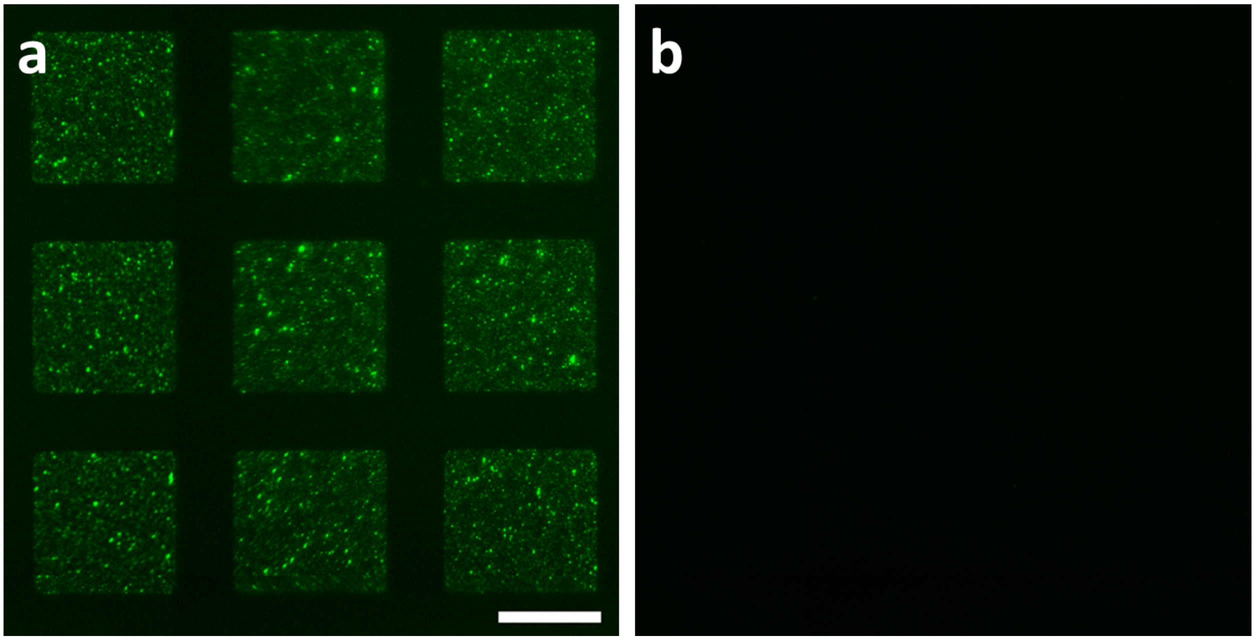


**Figure S2**: **DNA-directed patterning to verify oligonucleotide labeling of liposomes**. **a** DiO liposomes labeled with cholesterol-tagged oligonucleotides bind, via hybridization, to specific regions of a glass slide where the complementary oligonucleotide has been patterned, in this case an array of 141 µm by 141 µm squares. **b** There is no evidence of nonspecific binding of unlabeled DiO liposomes to DNA patterned arrays. Scale bar = 100 µm.

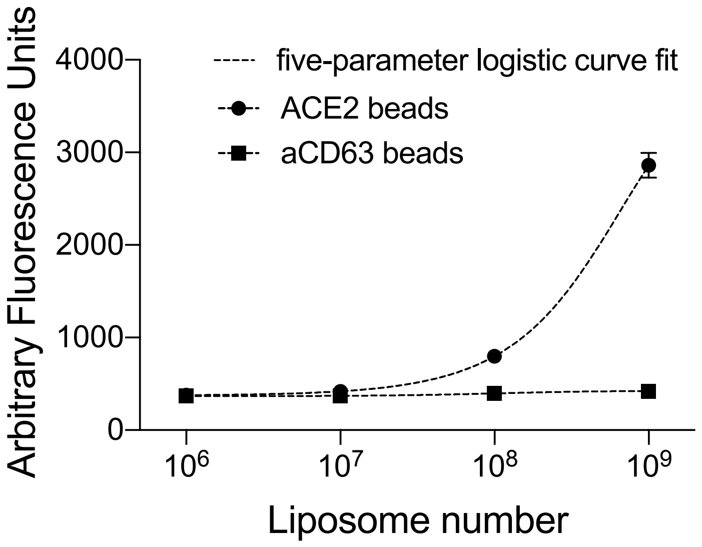


**Figure S3**: **Capture of fluorescent spike-liposomes onto ACE2 beads or aCD63 beads**. Mean fluorescent intensity was strongly correlated with spike-liposome number for ACE2 beads ($r^2 > 0.99$, $p = 0.002$), whereas for aCD63 beads, there was no significant correlation ($r^2 = 0.81$, $p = 0.097$). Pearson's correlation; error bars represent SEM. Dashed lines indicate least-squares five-parameter logistic curve fits ($r^2 = 0.91$ and 0.28 for ACE2 and aCD63 beads, respectively). For detailed statistical information, see Table S7.

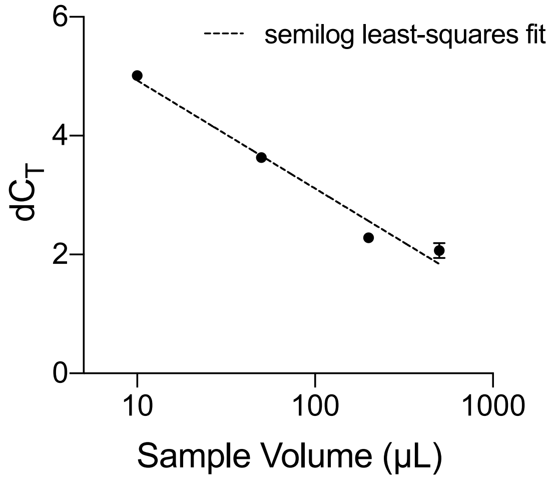


**Figure S4**: **qPCR signal as a function of sample volume (after sample processing).** A 50x increase in sample volume results in an 8-fold (3 cycle) decrease in signal, demonstrating that our method is more sensitive to the total amount, rather than the concentration, of target nanoparticles in a sample. Semilog least-squares fit: $r^2 = 0.97$; error bars indicate SEM.

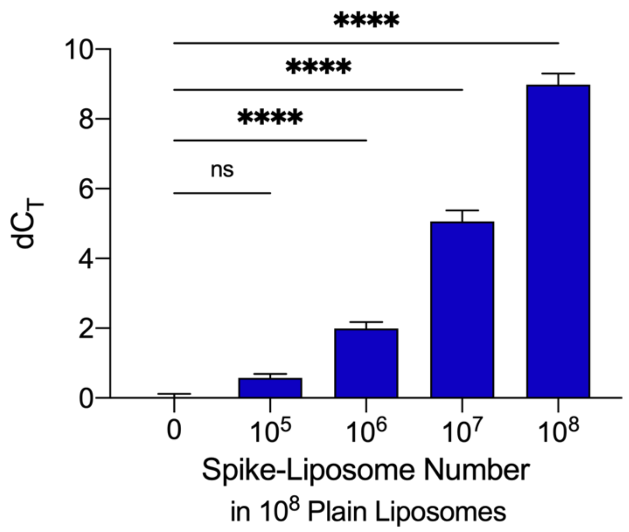


**Figure S5**: **Detection of 1x10$^6$ spike-liposomes in background of 1x10$^8$ plain liposomes**. We spiked a dilution series of spike-liposomes (1x10$^5$ – 1x10$^8$ particles) into a solution of 1x10$^8$ plain liposomes (20 µL total volume), incubated the resulting solution with 500 fmol of oligonucleotide label, and captured spike-liposomes onto 1x10$^6$ ACE2 beads. Error bars represent SEM; ns: not significant, ****: p<0.0001, one-way ANOVA with Dunnett's multiple comparisons test, with n = 6 independent replicates, each with n = 3 technical replicates. For detailed statistical information, see Table S8.

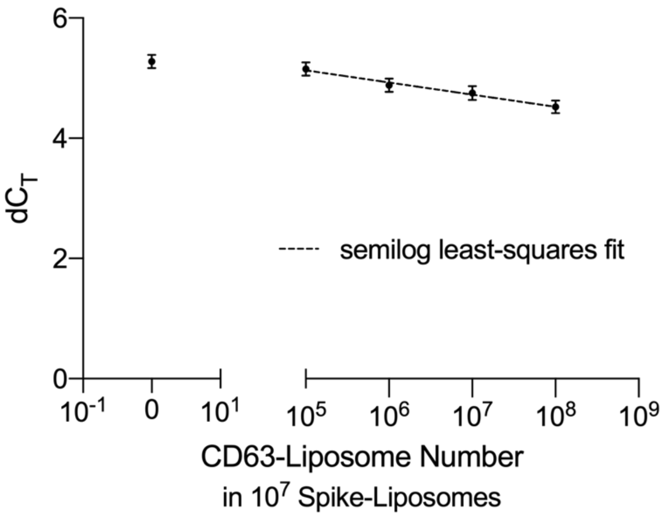


**Figure S6**: **qPCR signal from 1x10$^7$ spike-liposomes captured on ACE2 beads in a background of CD63 liposomes**. dCt, the difference between a sample containing liposomes and a no-liposome control, decreases by less than one cycle with the addition of 10$^8$ CD63-liposomes. This decrease is monotonic with an increasing number of background liposomes. Semilog least-squares fit: $r^2 = 0.67$; error bars indicate SEM.

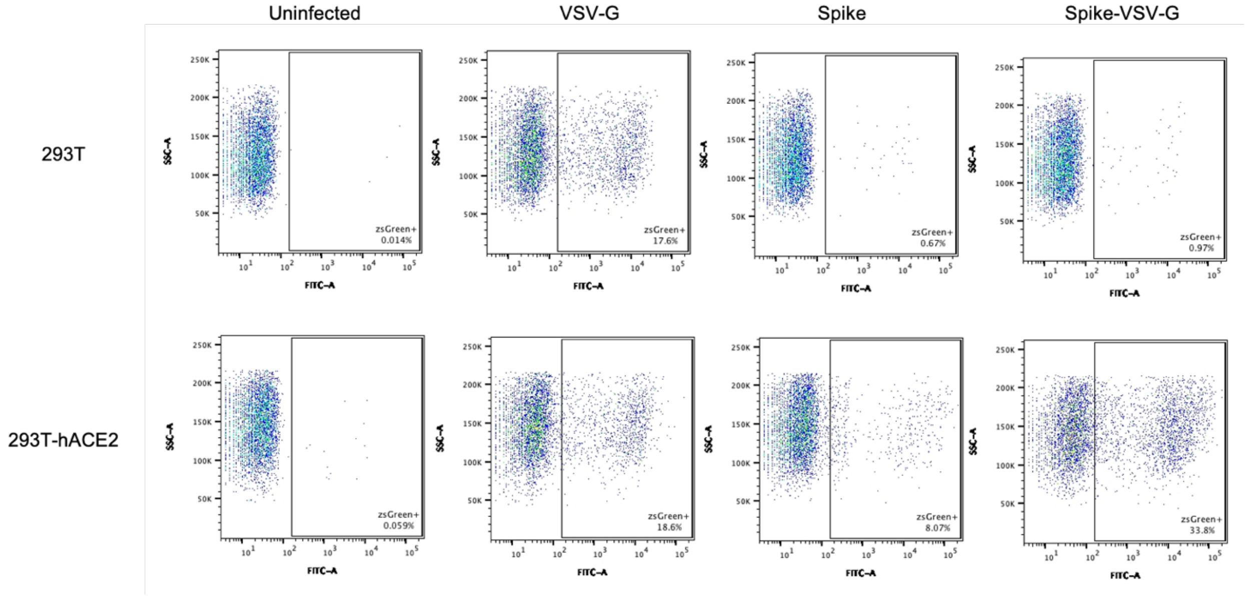


**Figure S7**: **Flow cytometry analysis showing percentage of cells infected with VSV-G, Spike and Spike-VSV-G chimera pseudotyped lentivirus.** Representative plots show side scatter versus ZsGreen, the fluorescent reporter found in the lentiviral backbone (FITC channel). For each virus, we selected conditions where 5-40% of the cells were infected (zsGreen$^+$ cells). Under the same conditions, infection of 293T-hACE2 cells with Spike-VSV-G resulted in a higher number of infected cells (33.8%) in comparison to WT Spike (8.07% infected cells).

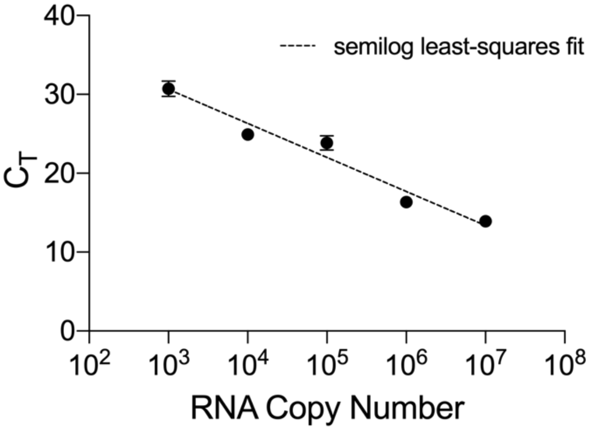


**Figure S8**: **Lenti-X standard curve.** This standard curve was used to calculate RNA copy number as a function of threshold cycle for spike- and VSV-G-pseudotyped lentivirus samples processed and analyzed in parallel. Semilog least-squares fit: $r^2 = 0.96$; error bars indicate SEM.

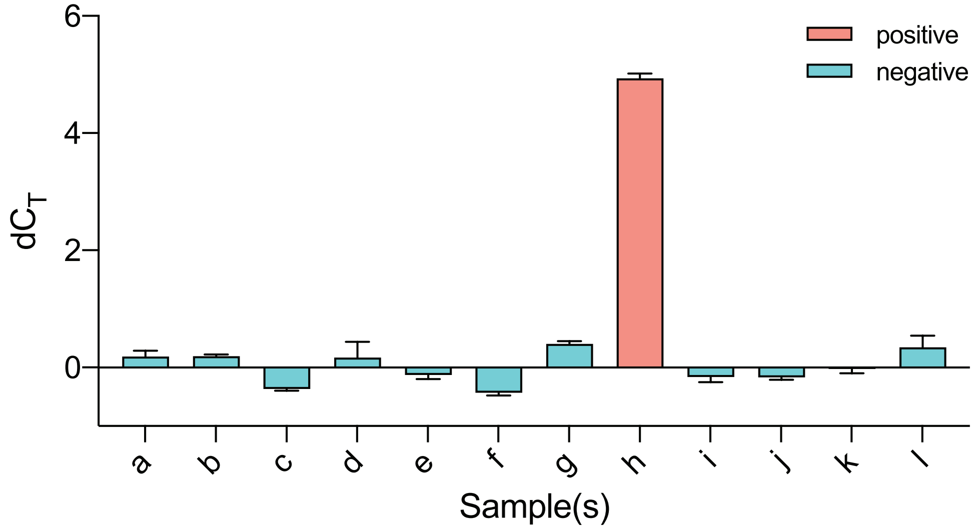


**Figure S9**: **qPCR signal for individual samples in spike-liposome sample pooling experiment.** All samples were processed, including those known to be negative based on an earlier pooling round, for confirmation. Negligible background signal was observed for all samples not containing spike liposomes (a-g, i-l). All samples contained $1x10^7$ CD63 liposomes per µL. Positive samples and pools, those containing at least one sample with spike liposomes ($1x10^6$/µL), produced a signal at least 8-fold higher than the threshold of 0.5 $dC_T$. n = 3 technical replicates; error bars indicate SEM.

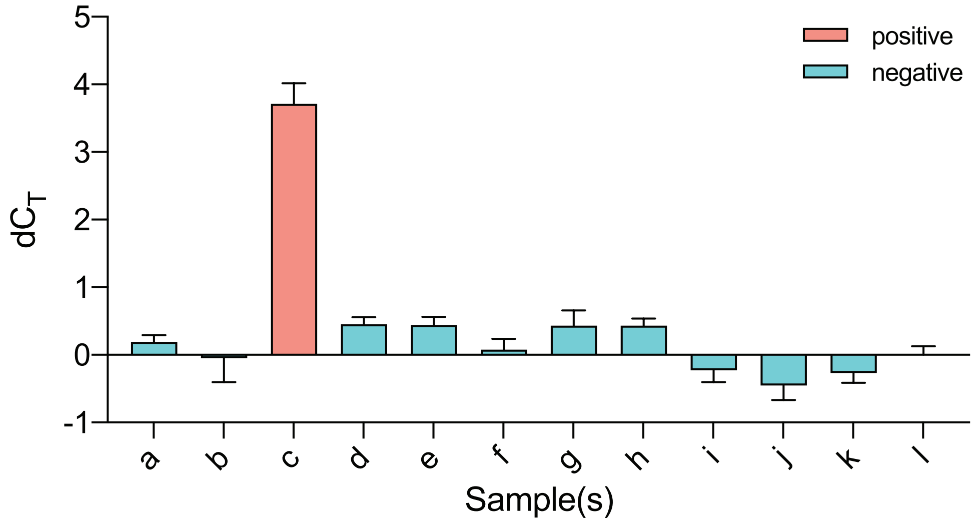


**Figure S10**: **qPCR signal for individual samples in Spike-pseudotyped lentivirus sample pooling experiment.** All samples were processed, including those known to be negative based on an earlier pooling round, for confirmation. Negligible background signal was observed for all samples not containing spike liposomes (a,b,d-l). Negative individual samples contained $4.02x10^6$/µL EVs, while positive individual samples contained $4.02x10^6$/µL EVs and 20 pfu/µL Spike-VSV-G-pseudotyped lentivirus. Positive samples and pools, those containing at least one sample with Spike-VSV-G lentivirus, produced a signal higher at least 1.5-fold higher than the threshold of 0.6 $dC_T$. n = 3 technical replicates; error bars indicate SEM.
